# Metabolic syndrome in Nepal: a systematic review and meta-analysis of prevalence, heterogeneity, and public health implications

**DOI:** 10.1101/2025.09.05.25335094

**Authors:** Sandip Pandey, Apil Tiwari, Alisha Bhattarai, Anu Timalsina, Asmit Pandey, Aashis Poudel, Abhikshya Basnet, Pramit Khatiwada, Aakash Neupane, Bibek Ojha

## Abstract

**Background:** Metabolic syndrome (MetS) is the clustering of abdominal obesity, dyslipidaemia, hypertension, and elevated fasting glucose. It raises the risk of cardiovascular disease and type 2 diabetes. In Nepal, reported MetS prevalence ranges from less than 5% to more than 50%. This variability reflects genuine population differences as well as methodological inconsistency. Pooled estimates and a systematic description of individual risk components are needed to guide national non-communicable disease policy.

**Methods:** We conducted a pre-registered systematic review and meta-analysis (PROSPERO CRD420251105131). We searched PubMed, Embase, Scopus, and four Nepalese journal portals from January 2009 to May 2025. Eligible studies enrolled Nepalese adults aged 15 years or older, reported MetS prevalence using the NCEP ATP III 2001 criteria, and included at least 100 participants. Risk of bias was assessed with the Joanna Briggs Institute (JBI) checklist. Certainty of evidence was rated using the GRADE framework. The primary analysis used a binomial generalised linear mixed model (GLMM) with a logit link. The 95% prediction interval (PI) was the main measure of dispersion. Subgroup analysis compared urban with suburban and mixed settings. A leave-one-out sensitivity analysis and Egger regression test for small-study effects were also performed.

**Results:** Eight cross-sectional studies enrolling 21,708 participants were included. The pooled MetS prevalence was 21.3% (95% CI: 11.6 to 35.9%). Between-study heterogeneity was extreme (I2 = 99.4%, tau2 = 1.08). The 95% PI ranged from 2.0% to 78.6%, meaning the true MetS prevalence in any given Nepalese community could be very low or very high. Urban and suburban or mixed settings did not differ significantly (20.0% versus 22.8%; p = 0.824). Among individual components, low HDL cholesterol was most prevalent (64.5%; 95% CI: 55.3 to 72.7%), followed by high waist circumference (59.5%; 95% CI: 34.8 to 80.1%) and elevated triglycerides (47.0%; 95% CI: 23.3 to 72.1%). Hypertension and elevated fasting glucose were present in 30.4% and 23.7% of participants, respectively. The leave-one-out analysis confirmed robustness of the pooled estimate (range: 18.0 to 26.2%). Egger regression did not detect significant small-study effects (p = [INSERT actual p-value from R output]). Three studies were rated low risk of bias and five moderate risk. GRADE certainty was very low for overall MetS prevalence, low for HDL cholesterol and hypertension, and very low for the remaining components.

## Background

Metabolic syndrome is defined as the co-occurrence of abdominal obesity, atherogenic dyslipidaemia, elevated blood pressure, and impaired fasting glucose. The syndrome confers a substantially higher risk of cardiovascular disease and type 2 diabetes than any single component alone (1). Global prevalence estimates range from 7% to over 40%. This variation reflects differences in population genetics, dietary patterns, physical activity, urbanisation, and the diagnostic criteria used (2).

In South Asia, rapid nutritional transition and increasingly sedentary lifestyles have driven a high and rising prevalence of MetS. In India, approximately one-third of urban adults in major cities meet the diagnostic criteria. Prevalence is notably higher in women than in men in several Indian studies (3), (4), (5).

Nepal presents a distinctive epidemiological setting. The country carries a dual burden of disease. Persistent undernutrition exists alongside growing cardiometabolic risk. Nepal’s geographical diversity spans the tropical Terai lowlands, the mid-hill zone, and Himalayan communities above 3,500 metres. Diet, physical activity, and access to healthcare differ substantially across these zones, and all three influence metabolic risk(6) (7). Communities in the Terai consume more refined carbohydrates and have higher rates of obesity and hypertension. High-altitude populations show distinct lipid profiles that may be partially explained by altitude-related physiology (8). This geographical diversity is compounded by ethnic heterogeneity across more than 125 caste and ethnic groups with differing dietary traditions and genetic backgrounds.

Available studies on MetS in Nepal report widely variable prevalence, from under 5% in young students to over 50% in older community-dwelling adults(9). Most studies are cross-sectional, regionally restricted, and inconsistent in reporting individual syndrome components. High-altitude communities and rural populations are under-represented. No previous systematic review has quantified the pooled prevalence of each individual MetS component alongside the overall syndrome. Component-level data are essential for prioritising interventions, particularly in settings where full-syndrome prevalence data are imprecise(10).

This systematic review and meta-analysis had four objectives. First, to estimate the pooled prevalence of MetS among Nepalese adults using the NCEP ATP III 2001 criteria. Second, to quantify the pooled prevalence of each individual MetS component. Third, to measure and explore sources of between-study heterogeneity. Fourth, to assess the certainty of the pooled evidence using the GRADE framework. These findings are intended to support targeted non-communicable disease interventions in Nepal and to identify methodological priorities for future research.

## Methods

### Protocol registration and reporting

The review protocol was registered with PROSPERO ([INSERT correct registration number, format CRD420251105131]). The original protocol was amended during the review to extend the search to May 2025 and to add sensitivity analyses for individual MetS components. This report follows the PRISMA 2020 guidelines(10). The completed PRISMA 2020 checklist is provided as Supporting Information (S1 Checklist).

### Eligibility criteria

Studies were eligible if they met all of the following five criteria.

- Enrolled Nepalese adults aged 15 years or older. The ATP III criteria were validated in adults aged 18 years and older; a pre-specified sensitivity analysis restricted to studies with a mean participant age of 18 years or older was performed to examine the effect of this boundary.
- Reported MetS prevalence using the NCEP ATP III 2001 diagnostic criteria(11). ATP III was chosen because it was the most consistently applied definition in the Nepali literature, enabling direct comparability across included studies. Studies using IDF, JIS, or WHO criteria were not eligible for the primary analysis. This restriction is acknowledged as a limitation and is discussed below.
- Observational in design: cross-sectional survey, cohort study at baseline, or nationally representative survey.
- Provided sufficient data to extract the number of MetS cases and the total sample size.
- Enrolled at least 100 participants.

Studies were excluded if participants were pre-selected on the basis of an existing high-risk condition, such as central obesity or non-alcoholic fatty liver disease, because such designs inflate the observed MetS prevalence. Case reports, conference abstracts, reviews, and meta-analyses were also excluded. Studies conducted outside Nepal were not eligible. Full-text review was limited to English-language sources; no formal exclusion of non-English records was applied during the database search stage.

### Information sources and search strategy

We searched PubMed, Embase, Scopus, the Journal of Nepal Medical Association, the Nepalese Heart Journal, and Nepal Journals Online. The search covered 1 January 2009 to 1 May 2025. The start date was chosen because no Nepal-specific MetS prevalence study meeting our inclusion criteria appeared before 2009 in preliminary searches. The search combined controlled vocabulary and free-text terms as follows:

("metabolic syndrome" OR "insulin resistance" OR "MetSyn") AND ("prevalence" OR "epidemiology") AND ("Nepal" OR "Himalaya*") NOT ("case report" OR "case series" OR "meta-analysis" OR "systematic review")

The full search strategy for each database is provided in Supporting Information (S2 Text). Search results were imported into Rayyan and de-duplicated. Two reviewers (SP, AT) independently screened titles and abstracts. A third reviewer (AB) resolved disagreements. Full texts were assessed against the eligibility criteria by the same pair of reviewers.

Unpublished studies were not retrieved.

### Data Extraction and Quality Assessment

Two reviewers (AT, AnT) independently extracted data using a standardised form. Extracted variables included study region and setting, year of data collection, study design, participant age range and mean, sex distribution where reported, diagnostic criteria, total sample size, number and percentage with MetS, and prevalence of each of the five ATP III components: waist circumference, HDL cholesterol, triglycerides, blood pressure, and fasting glucose. Where a study reported prevalence as a percentage only, event counts were back-calculated from the sample size. Sex-stratified prevalence was extracted wherever reported. Where studies did not provide sex-disaggregated data, this was recorded as not reported (NR). Disagreements were resolved by consensus or by a third reviewer.

Methodological quality was assessed using the JBI Critical Appraisal Checklist for Studies Reporting Prevalence Data. This tool covers nine domains. Each domain was rated Yes, No, or Unclear. Studies were classified as low risk (seven or more Yes ratings), moderate risk (four to six Yes ratings), or high risk (three or fewer Yes ratings). The full domain-level assessment is provided in Supporting Information (S2 Text); a summary is presented in Table 4.

### Subgroup classification

Studies were classified as urban if recruitment took place in the Kathmandu Valley or if the authors explicitly described the setting as city-based (14). Studies recruiting from community-based samples outside the Kathmandu Valley, or combining urban and peri-urban or rural catchments, were classified as suburban or mixed. Two reviewers classified studies independently and resolved discrepancies by consensus.

### Statistical Analysis

For each study, the proportion and exact binomial 95% confidence interval were calculated. The primary meta-analysis used a GLMM with a binomial distribution and logit link, estimated by maximum likelihood (12). This model is appropriate for pooling proportions because it does not require a transformation of raw data and is robust when event rates are very low or very high. It avoids the bias that can affect the DerSimonian-Laird estimator in these situations.

A pre-specified sensitivity analysis used the DerSimonian-Laird random-effects model with logit-transformed proportions and the Hartung-Knapp adjustment for confidence intervals, estimated by restricted maximum likelihood (REML) (13). This provided a methodologically independent check on the primary estimate.

Heterogeneity was quantified using I2 and tau2. Given the extreme between-study heterogeneity observed (I2 above 99%), the 95% prediction interval is emphasised throughout as the primary measure of dispersion. The prediction interval reflects the expected range of the true effect in a new population drawn from the same distribution of study contexts [17]. Subgroup analysis compared urban with suburban or mixed settings. There were insufficient exclusively rural studies to form a separate subgroup.

Separate meta-analyses were conducted for each of the five ATP III components. Leave-one-out meta-analysis and cumulative meta-analysis ordered by year of publication were performed to assess robustness and to detect time trends.

Small-study effects and possible publication bias were assessed using Egger regression applied to logit-transformed proportions [18]. With eight studies, statistical power to detect funnel plot asymmetry is low. The test is therefore reported as a supplementary diagnostic rather than definitive evidence for or against publication bias. The funnel plot is provided in Supporting Information (S3 Fig).

All analyses were performed in R version 4.3.0 using the meta package (version 6.5-0) and the metafor package (version 4.4-0) [18, 19]. The analysis code and extracted data are publicly available at https://github.com/Dr-Charlie/metabolic-syndrome-meta-analysis.

### Certainty of Evidence Assessment

The certainty of evidence for each pooled estimate was rated using the GRADE approach (14). Ratings were assigned independently by two reviewers (BO, PK) and disagreements were resolved by consensus. The full GRADE summary table is provided in Supporting Information (S3 Text); a summary is presented in Table 5.

## Results

### Study Selection

Database searches identified 115 records. Screening of journal websites identified two additional records. After removing 55 duplicates and five records excluded for other reasons, 55 records were screened by title and abstract. Forty-four were excluded at this stage. Of 11 full-text reports sought, two could not be retrieved. Nine full-text reports were assessed for eligibility. Three were excluded: one enrolled only participants with central obesity, and two enrolled only participants with non-alcoholic fatty liver disease. Two additional reports from local journal sources were both eligible. In total, eight studies met the inclusion criteria and were included in the qualitative synthesis and the meta-analysis. The PRISMA 2020 flow diagram is presented as Figure 1. Excluded studies and reasons are listed in Table 1.

**Figure 1.** [PRISMA Flowdiagram]

**Table 1.**
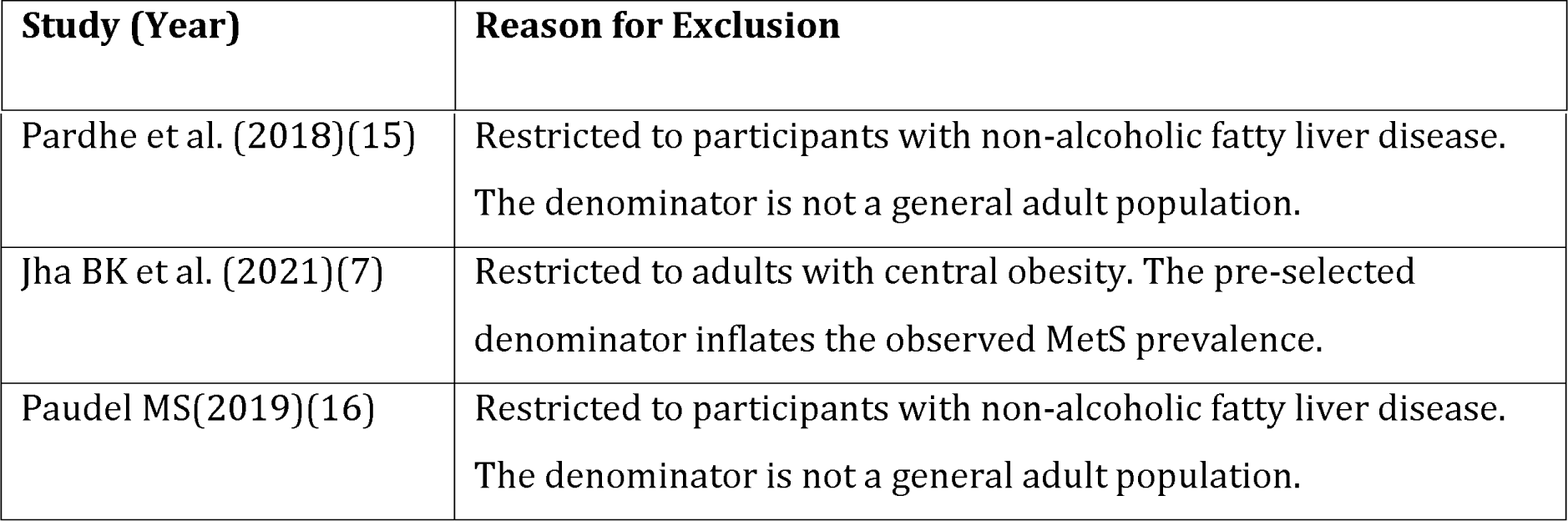
Studies excluded at full-text review and reasons for exclusion.

| Study (Year) | Reason for Exclusion |
| --- | --- |
| Pardhe et al. (2018)(15) | Restricted to participants with non-alcoholic fatty liver disease.<br>The denominator is not a general adult population. |
| Jha BK et al. (2021)(7) | Restricted to adults with central obesity. The pre-selected denominator inflates the observed MetS prevalence. |
| Paudel MS(2019)(16) | Restricted to participants with non-alcoholic fatty liver disease.<br>The denominator is not a general adult population. |

### Study Characteristics

All eight included studies were cross-sectional in design. They were published between 2009 and 2025 and enrolled a total of 21,708 participants. Sample sizes ranged from 145 to 14,425 participants. Participant ages spanned 15 to 86 years. Four studies recruited in urban Kathmandu settings, including hospital health check-up programmes and a student population. Four studies recruited from broader mixed or national samples. All eight studies applied the NCEP ATP III 2001 diagnostic criteria.

Individual MetS component data were incompletely reported across studies. All eight studies reported waist circumference and HDL cholesterol data. Triglyceride, blood pressure, and fasting glucose data were each available from only a subset of studies. NR is indicated where data were not reported. Sex-stratified MetS prevalence was not consistently reported across the included studies. Where individual studies provided sex-disaggregated data, this is noted in the text. Characteristics of all included studies are presented in Table 2.

**Table 2.**
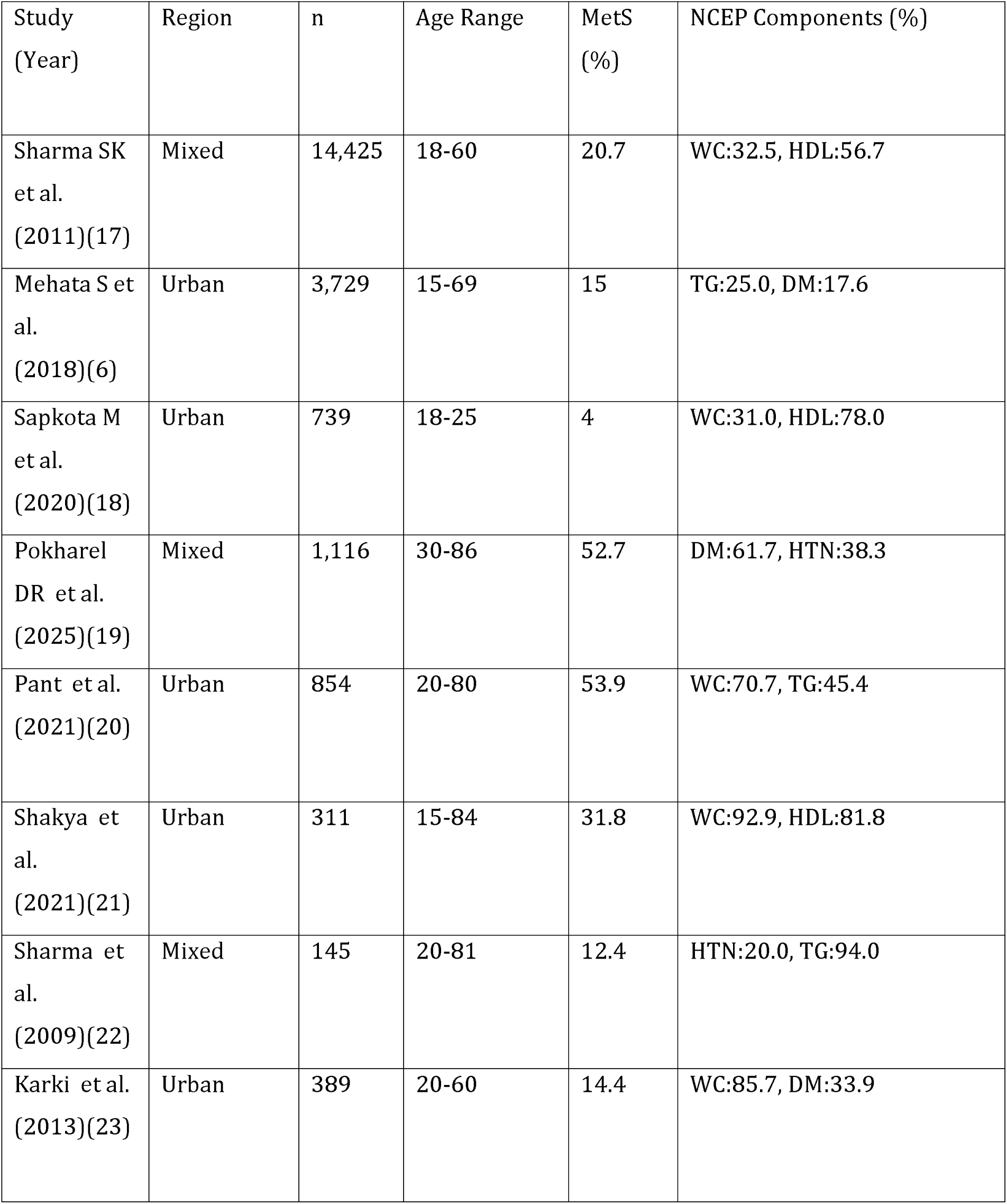
Characteristics of included studies and prevalence of individual metabolic syndrome components (NCEP ATP III 2001). *WC = waist circumference; HDL = high-density lipoprotein cholesterol; TG = triglycerides; HTN = hypertension; DM/HG = diabetes or elevated fasting glucose; NR = not reported. MetS prevalence is reported as % (n)*.

| Study (Year) | Region | n | Age Range | MetS (%) | NCEP Components (%) |
| --- | --- | --- | --- | --- | --- |
| Sharma SK et al. (2011)(17) | Mixed | 14,425 | 18-60 | 20.7 | WC:32.5, HDL:56.7 |
| Mehata S et al. (2018)(6) | Urban | 3,729 | 15-69 | 15 | TG:25.0, DM:17.6 |
| Sapkota M et al. (2020)(18) | Urban | 739 | 18-25 | 4 | WC:31.0, HDL:78.0 |
| Pokharel DR et al. (2025)(19) | Mixed | 1,116 | 30-86 | 52.7 | DM:61.7, HTN:38.3 |
| Pant et al. (2021)(20) | Urban | 854 | 20-80 | 53.9 | WC:70.7, TG:45.4 |
| Shakya et al. (2021)(21) | Urban | 311 | 15-84 | 31.8 | WC:92.9, HDL:81.8 |
| Sharma et al. (2009)(22) | Mixed | 145 | 20-81 | 12.4 | HTN:20.0, TG:94.0 |
| Karki et al. (2013)(23) | Urban | 389 | 20-60 | 14.4 | WC:85.7, DM:33.9 |

### Overall pooled prevalence

The pooled MetS prevalence from the primary GLMM model was 21.3% (95% CI: 11.6 to 35.9%). The sensitivity analysis using the DerSimonian-Laird REML model with Hartung-Knapp adjustment yielded a similar estimate of 21.5% (95% CI: 9.8 to 40.9%). Between-study heterogeneity was extreme (I2 = 99.4%, tau2 = 1.08, p < 0.0001). The forest plot is presented as Figure 2.

**Figure 2.** [insert Overall Pooled Prevalence of Metabolic Syndrome]

### Prediction interval and heterogeneity

The 95% prediction interval ranged from 2.0% to 78.6%. This is the most informative result of the analysis. It indicates that the true MetS prevalence in a new Nepalese community could lie anywhere within this wide range. A national point estimate of approximately 21% must not be interpreted as representative of any specific community or population subgroup.

### Subgroup analysis by study setting

The pooled prevalence was 20.0% (95% CI: 6.9 to 45.7%) in urban studies and 22.8% (95% CI: 11.8 to 39.5%) in suburban or mixed studies. The test for subgroup differences was not significant under the random-effects model (p = 0.824). Geographic setting did not explain the observed heterogeneity. Substantial residual heterogeneity remained within each subgroup (I2 = 99.2% and 99.5%, respectively). The subgroup forest plot is presented as Figure 3.

**Figure 3.** [Insert subgroup analysis by site]

### Prevalence of individual components

Separate meta-analyses were performed for each of the five NCEP ATP III components. Low HDL cholesterol was the most prevalent component, with a pooled prevalence of 64.5% (95% CI: 55.3 to 72.7%). High waist circumference was found in 59.5% of participants (95% CI: 34.8 to 80.1%). Elevated triglycerides were present in 47.0% (95% CI: 23.3 to 72.1%). Hypertension was found in 30.4% (95% CI: 18.0 to 46.4%) and elevated fasting glucose in 23.7% (95% CI: 11.3 to 43.2%). All component analyses showed high heterogeneity (I^2^ above 98%). Forest plots for HDL cholesterol and waist circumference are presented as Figures 4a and 4b. Forest plots for the remaining three components are provided in Supporting Information (S4 Fig to S6 Fig). These component-level findings are clinically important. The pooled prevalence of low HDL (64.5%) and abdominal obesity (59.5%) is far higher than the full-syndrome prevalence (21.3%). This means that a large proportion of the Nepalese adult population carries one or more high-risk MetS components without meeting the full diagnostic threshold. These individuals face elevated cardiovascular risk that warrants clinical attention.

**Figure 4a:** [Insert Forest Plots of MetS Component Prevalence(low HDL)]

**Figure 4b:** [Insert Forest Plots of MetS Component Prevalence(Waist circumference)]

Consistent sex-stratified data for individual components were not available from a sufficient number of included studies to permit formal meta-analysis by sex. This represents a significant evidence gap, given the well-documented sex differential in MetS prevalence across South Asia.

### Sensitivity and Influence Analyses

The leave-one-out analysis confirmed robustness of the pooled estimate. After sequentially omitting each study, the recalculated prevalence ranged from 18.0% to 26.2%. All estimates fell within the 95% CI of the primary analysis (11.6 to 35.9%). No single study exerted undue influence on the overall result. Results are presented in Table 3. The leave-one-out forest plot is presented as Figure 5.

**Figure 5.** [Insert Leave one out analysis]

**Table 3.**
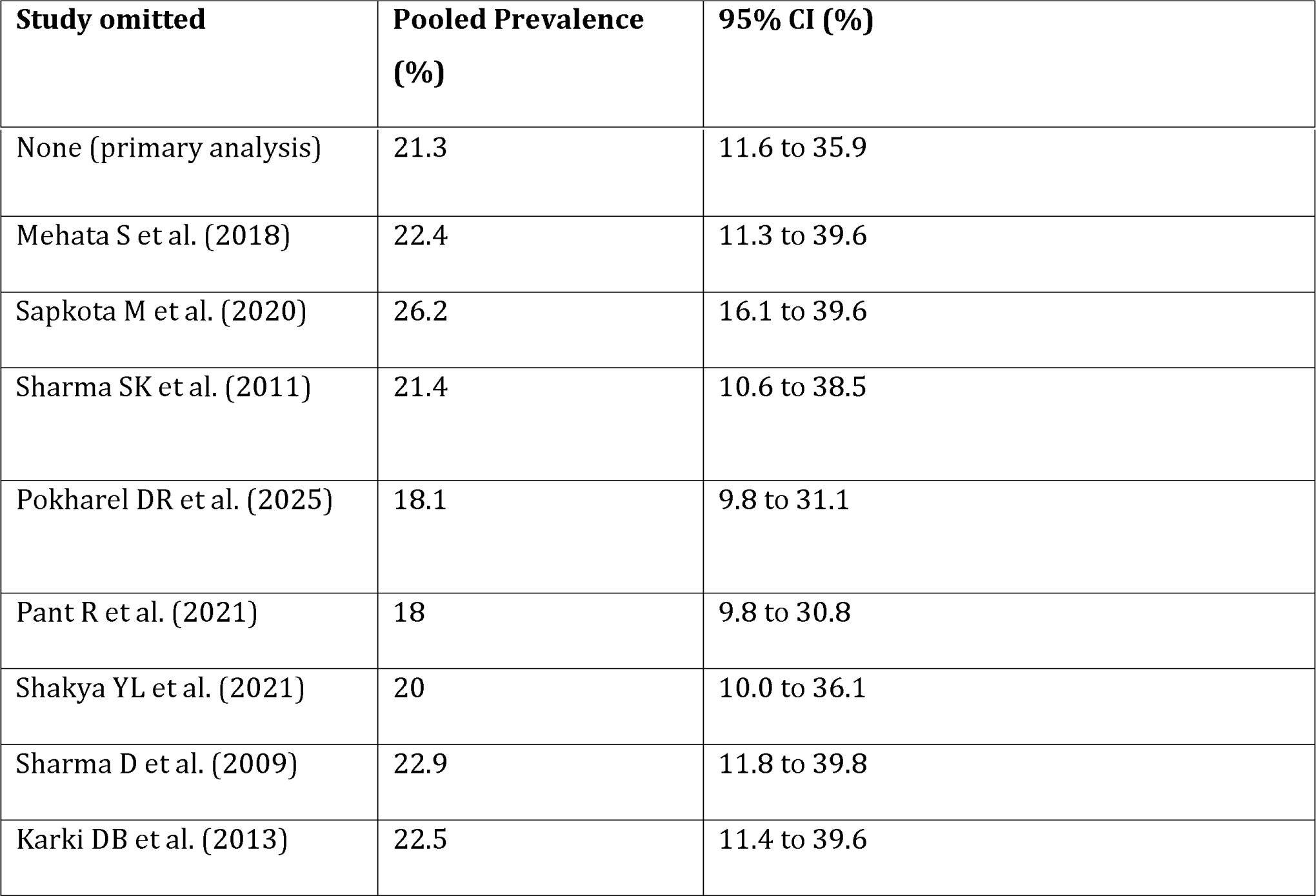
Leave-one-out sensitivity analysis.

| Study omitted | Pooled Prevalence (%) | 95% CI (%) |
| --- | --- | --- |
| None (primary analysis) | 21.3 | 11.6 to 35.9 |
| Mehata S et al. (2018) | 22.4 | 11.3 to 39.6 |
| Sapkota M et al. (2020) | 26.2 | 16.1 to 39.6 |
| Sharma SK et al. (2011) | 21.4 | 10.6 to 38.5 |
| Pokharel DR et al. (2025) | 18.1 | 9.8 to 31.1 |
| Pant R et al. (2021) | 18 | 9.8 to 30.8 |
| Shakya YL et al. (2021) | 20 | 10.0 to 36.1 |
| Sharma D et al. (2009) | 22.9 | 11.8 to 39.8 |
| Karki DB et al. (2013) | 22.5 | 11.4 to 39.6 |

The Baujat plot (Figure 6) identified two studies as the largest contributors to overall heterogeneity. Sharma SK et al. (2011) was the largest study (n = 14,425) and used a national sample. Pokharel DR et al. (2025) enrolled an older, community-dwelling sample aged 30 to 86 years (n = 1,116). These differences in study population likely reflect genuine variation in MetS burden rather than methodological error.

**Figure 6.** [Insert Baujat plot]

The cumulative meta-analysis (Figure 7) showed that the pooled estimate was unstable when only the first two or three studies were available. As subsequent studies were added, the estimate stabilised within a range of 16% to 21%. The final estimate of 21.3% reflects a converged body of evidence rather than being driven by early studies.

**Figure 7.** [Insert Cumulative Meta-Analysis]

### Publication bias and small-study effects

Egger regression applied to logit-transformed proportions did not detect statistically significant small-study effects (intercept = [INSERT value]; 95% CI: [INSERT CI]; p = [INSERT p-value]). With only eight studies, statistical power to detect moderate publication bias is limited. The funnel plot (Supporting Information S3 Fig) shows some asymmetry, with smaller studies tending towards higher prevalence estimates. This pattern is consistent with both publication bias and genuine population heterogeneity. A definitive conclusion cannot be drawn from these data alone.

### Risk of Bias Assessment

The JBI domain-level risk of bias assessment is presented in Table 4. All eight studies drew on appropriate sample frames and applied the NCEP ATP III criteria consistently, yielding Yes ratings for domains 1 and 6 across all studies. Six studies used random or stratified sampling. Two studies relied on convenience sampling, which raises concerns about representativeness. Three studies were judged to have inadequate sample sizes. Response rates were explicitly reported in only three studies; the remaining five received Unclear ratings for domain 9. Three studies were rated low risk of bias overall and five were rated moderate risk. No study was rated high risk.

**Table 4.** JBI Critical Appraisal Checklist: domain-level risk of bias assessment. Y = Yes; N = No; U = Unclear.

### Certainty of evidence (GRADE)

GRADE ratings for all six outcomes are presented in Table 5. The certainty of evidence for overall MetS prevalence was rated very low. This rating reflects serious risk of bias, very serious inconsistency (I2 = 99.4%), and serious imprecision. Low HDL cholesterol and hypertension received low certainty ratings. These two outcomes were downgraded for risk of bias and very serious inconsistency but not for imprecision, as their confidence intervals were comparatively narrow. All remaining component estimates received very low certainty ratings, primarily because of extreme heterogeneity and imprecision. No outcome was downgraded for indirectness, as the population and diagnostic criteria were consistent with the review question throughout.

**Table 5.** GRADE certainty of evidence summary.

## Discussion

### Principal findings

This meta-analysis of eight studies enrolling 21,708 Nepalese adults produced three main findings. First, the pooled MetS prevalence of 21.3% (95% CI: 11.6 to 35.9%) is consistent with mid-range South Asian estimates, which commonly fall between 14% and 33%(24). Second, the 95% prediction interval of 2.0% to 78.6% reveals extreme heterogeneity. The pooled mean is misleading as a policy target. The true MetS burden in any specific Nepalese community may be far lower or far higher than the national average. Third, the pooled prevalences of low HDL cholesterol (64.5%) and abdominal obesity (59.5%) are high, consistent across settings, and carry independent clinical significance. These component-level findings are the most actionable outputs of the review (25).

### Interpretation of heterogeneity

Extreme heterogeneity in prevalence meta-analyses is common when studies span diverse populations, settings, and time periods. Several factors that could not be formally tested in meta-regression plausibly explain the observed variation in this review. The number of included studies was insufficient for a reliable meta-regression analysis.

Age composition is a likely contributor. Studies ranged from young health-sciences students aged 18 to 25 years, with a MetS prevalence of 4.1% (Sapkota et al., 2020), to a community sample aged 30 to 86 years with a prevalence of 52.7% (Pokharel et al., 2025). MetS prevalence rises steeply with age. Selection mechanism is another factor. Hospital-based health check-up samples are enriched for individuals with pre-existing cardiometabolic concerns, which inflates observed prevalence (Pant et al., 53.9%; Shakya et al., 31.8%). Ecological zone and ethnicity also differ across studies, but no study provided sufficient data to classify participants by altitude or ethnic group. Finally, the 16-year span of included publications (2009 to 2025) encompasses substantial nutritional transition in Nepal. The cumulative analysis suggests a possible secular increase in prevalence, though this cannot be confirmed with the available data.

The failure of the urban versus suburban subgroup analysis to explain heterogeneity (p = 0.824) confirms that a coarse geographic binary is an inadequate proxy for the underlying drivers of MetS risk. The Baujat plot identified Sharma SK et al. (2011) and Pokharel DR et al. (2025) as the largest contributors to heterogeneity. These two studies represent the largest nationally representative survey and the oldest community sample in the dataset. Their divergent prevalence estimates reflect genuine population-level differences rather than methodological problems warranting exclusion.

### Component-level burden and clinical significance

The high prevalence of low HDL (64.5%) and abdominal obesity (59.5%) has greater certainty than the overall MetS estimate and is directly actionable. Both components respond to the same lifestyle interventions: reduced intake of refined carbohydrates and saturated fats, increased consumption of vegetables, legumes, and unsaturated fats, and increased physical activity. The South Asian lipid phenotype, characterised by low HDL and high triglycerides with only mildly elevated LDL, is recognised as particularly atherogenic. It may explain why cardiovascular risk in South Asians is higher than conventional risk scores predict (26),(27).

A key clinical implication is that component-level screening and management are worthwhile even when full MetS criteria are not met. An individual with two components who does not satisfy the full diagnostic threshold still faces elevated cardiovascular and diabetes risk. Early intervention at the component level can reduce this risk before it progresses to a full metabolic syndrome.

### Comparison with regional evidence

Nepal’s pooled estimate of 21.3% falls within the range reported in neighbouring South Asian countries. Estimates from India, Bangladesh, and Sri Lanka commonly range from 18% to 33% depending on the region and criteria applied [21, 24]. Nepal shows markedly wider dispersion across studies than is typically seen in single-country reviews from India. This wider dispersion likely reflects the country’s exceptional ecological and ethnic diversity, compressed into a geographic area smaller than many Indian states. This internal heterogeneity is itself an important finding for health equity. Communities at the upper end of the prediction interval may carry a MetS burden comparable to that of high-income countries. Those at the lower end risk under-screening and under-treatment.

### Strengths and limitations

This review has several strengths. The protocol was prospectively registered with PROSPERO. Reporting follows the PRISMA 2020 statement. The primary analysis used a GLMM with binomial-logit link, which is methodologically superior to the DerSimonian-Laird estimator for proportion meta-analyses. The prediction interval was used as the primary dispersion metric. Extensive sensitivity analyses were performed, including leave-one-out analysis, cumulative meta-analysis, an alternative statistical model, and Egger regression. GRADE assessments were completed for all six outcomes.

The review also has important limitations. First, only eight studies met the eligibility criteria. The low yield of eight studies from 115 records reflects the restriction to NCEP ATP III 2001 as the sole eligible diagnostic criteria. This decision maximised comparability across included studies, as ATP III was the most consistently applied definition in the Nepali literature. However, it excludes studies that used the IDF or JIS criteria. Readers should interpret the pooled estimate as specific to ATP III-defined MetS and should not assume equivalence with estimates from other diagnostic frameworks. Second, all included studies are cross-sectional. Causal inference and temporal tracking of MetS trends are not possible from this evidence base. Third, sex-stratified data were not consistently reported. A formal meta-analysis by sex was not possible, despite the well-documented sex differential in South Asian MetS prevalence. Fourth, high-altitude communities, indigenous populations, and rural populations outside the Terai and Kathmandu Valley are under-represented in the evidence base. Fifth, two studies used convenience sampling and five did not report response rates, contributing to the risk of bias concerns noted in the quality assessment. Sixth, the full-text review was limited to English-language sources, which may have led to the exclusion of relevant Nepali-language material. Seventh, Egger regression has low power with fewer than ten studies. The absence of a significant result should not be interpreted as confirming the absence of publication bias.

### Implications for policy and practice

National averages should not drive programme design given the extreme heterogeneity across communities. Screening and intervention resources should be prioritised using district-level or municipality-level data rather than a national point estimate. Where local data are unavailable, communities with older age profiles, hospital catchment areas, Terai lowland locations, or sedentary occupational patterns should be prioritised for surveillance.

The consistent burden of dyslipidaemia and central adiposity across all settings justifies immediate nationwide action. Waist circumference measurement, fasting lipid profiles, and blood pressure screening should be integrated into existing primary care platforms, including the STEPS survey infrastructure and community health outreach programmes. Dietary guidance specific to the South Asian lipid phenotype should be incorporated into health education at community level.

For clinical practice, these findings support proactive metabolic screening for Nepalese adults, including younger adults and those without obvious risk factors. Component-based management can reduce downstream cardiovascular and diabetes risk before full MetS criteria are met.

### Priorities for future research

Future studies should use representative, stratified sampling across Nepal’s three ecological zones (Terai, Hills, and Mountains) and all major ethnic groups. Pre-specified reporting of sex-stratified data should be standard. Standardised diagnostic criteria, ideally reporting both ATP III and IDF results, would facilitate future meta-analyses. High-altitude communities and indigenous populations must be included. Longitudinal designs are needed to establish MetS trajectories and causal relationships with cardiovascular outcomes. For future synthesis, investigators should plan a priori for high inconsistency, emphasise prediction intervals, and pursue individual participant data meta-analysis where feasible.

## Conclusions

MetS affects approximately one in five Nepalese adults on average. This figure conceals extreme variation across communities and cannot serve as a reliable policy target. The most consistent and actionable findings are the high pooled burdens of low HDL cholesterol (64.5%) and abdominal obesity (59.5%). These two components independently raise cardiometabolic risk and respond to the same lifestyle interventions. Public health programmes should use localised screening data, address dyslipidaemia and central adiposity through diet and physical activity initiatives, and integrate metabolic screening into primary care. A coordinated programme of population-representative, sex-stratified, and ecologically diverse research is required to explain the heterogeneity that current evidence cannot resolve.

## Data Availability

All data produced in the present study are available upon reasonable request to the authors

https://github.com/Dr-Charlie/metabolic-syndrome-meta-analysis

## List of abbreviations

ATP III: Adult Treatment Panel III
CI: Confidence interval
GLMM: Generalised linear mixed model
GRADE: Grading of Recommendations Assessment, Development and Evaluation
HDL: High-density lipoprotein cholesterol
I2: Inconsistency statistic
IDF: International Diabetes Federation
JBI: Joanna Briggs Institute
MetS: Metabolic syndrome
ML: Maximum likelihood
NAFLD: Non-alcoholic fatty liver disease
NCEP: National Cholesterol Education Program
NR: Not reported
PI: Prediction interval
PRISMA: Preferred Reporting Items for Systematic Reviews and Meta-Analyses
PROSPERO: International Prospective Register of Systematic Reviews
REML: Restricted maximum likelihood
WC: Waist circumference

## Declarations

### Ethics approval and consent to participate

Not applicable. This study is a systematic review and meta-analysis of previously published literature. No new primary data were collected from human participants.

### Consent for publication

Not applicable.

### Availability of data and materials

All data generated or analysed during this study are included in this article and its Supporting Information files. The R analysis code and extracted data are available at https://github.com/Dr-Charlie/metabolic-syndrome-meta-analysis.

### Authors’ contributions

SP conceived the study, served as guarantor and corresponding author, supervised methods and analyses, and led drafting and revision. AP (Asmit Pandey) performed database searches, curated data, implemented software and figures, and contributed to drafting. AT refined methodology, provided statistical support and validation, and critically reviewed the manuscript. AB and AnT screened records, extracted data, contributed to risk of bias assessment, and revised the manuscript. ABas retrieved full texts, searched local sources, verified data, and reviewed the manuscript. BO led quality appraisal and GRADE assessment and assisted in interpretation. PK contributed to quality appraisal, clinical interpretation, and the policy implications section. AN provided health-system context, edited the manuscript, and supported data visualisation. All authors had access to the data, approved the final manuscript, and accept accountability for the work.

## Acknowledgements

The authors thank the investigators of all primary studies whose data contributed to this meta-analysis.

